# Combining genomic data and infection estimates to characterize the complex dynamics of SARS-CoV-2 Omicron variants in the United States

**DOI:** 10.1101/2023.11.07.23298178

**Authors:** Rafael Lopes, Kien Pham, Fayette Klaassen, Melanie H. Chitwood, Anne M. Hahn, Seth Redmond, Nicole A. Swartwood, Joshua A. Salomon, Nicolas A. Menzies, Ted Cohen, Nathan D. Grubaugh

## Abstract

SARS-CoV-2 Omicron surged as a variant of concern in late 2021. Subsequently, several distinct Omicron variants have appeared and overtaken each other. We combined variant frequencies and infection estimates from a nowcasting model for each US state to estimate variant-specific infections, attack rates, and effective reproduction numbers (R_t_). BA.1 rapidly emerged, and we estimate that it infected 47.7% of the US population between late 2021 and early 2022 before it was replaced by BA.2. We estimate that BA.5, despite a slower takeoff than BA.1, infected 35.7% of the US population, persisting in circulation for nearly 6 months. Other Omicron variants - BA.2, BA.4, and XBB - together infected 30.7% of the US population. We found a positive correlation between the state-level BA.1 attack rate and social vulnerability and a negative correlation between the BA.1 and BA.2 attack rates. Our findings illustrate the complex interplay between viral evolution, population susceptibility, and social factors during the Omicron emergence in the US.

## Introduction

Nearly four years since the World Health Organization declared the COVID-19 outbreak as a pandemic, SARS-CoV-2 caused more than 778 million confirmed cases globally and more than 6.9 million deaths (1). The emergence of genetically distinct SARS-CoV-2 variants of concern (VOC) posed a major challenge for control programs and greatly extended the length and health impact of the pandemic.

Following the emergence of the first major VOC, Alpha, in late 2020 (2), new VOCs have arisen and resulted in successive waves of infection (3,4). Alpha co-circulated with both Beta and Gamma variants (first detected contemporaneously in late 2020 in South Africa and Brazil (5,6), respectively); these variants were subsequently replaced after the emergence and spread of the Delta variant (7) in mid-2021. The emergence of the Omicron variant, first detected in South Africa and Botswana in November 2021 (8,9) was followed by rapid global spread and the replacement of the Delta variant.

Large-scale genomic sequencing of SARS-CoV-2 isolates collected from individuals with detected COVID-19 disease has been instrumental in documenting the evolution of successive VOC in many settings (5,7,9–11). However, a considerable fraction of SARS-CoV-2 infections do not result in documented disease (12–15), especially after the introduction of vaccines and the development of partial immunity associated with previous infection (16–18). Understanding the dynamics of transmission and strain replacement requires methods to infer time trends in variant-specific infections. Here, we combine nationwide SARS-CoV-2 sequencing data from GISAID with infection estimates from a Bayesian nowcasting model to better characterize the rise and fall of Omicron variants in the United States (US) between late 2021 and March 2023.

## Results

### Quantifying variant-specific infections by combining variant frequency and infection estimates

The emergence and spread of multiple SARS-CoV-2 variants has been a hallmark of the COVID-19 pandemic. Combining 3,103,250 SARS-CoV-2 genomic sequences (**Figs. S1-S3**) and infection estimates from a nowcasting model (*covidestim* (19); **Fig. 1A**), we estimated daily infections by each major variant of Omicron (BA.1*, BA.2*, BA.4*, BA.5*, and XBB*) from each US state and the District of Columbia from December 2021 to March 2023 (**Fig. 1**).

**Fig. 1.**
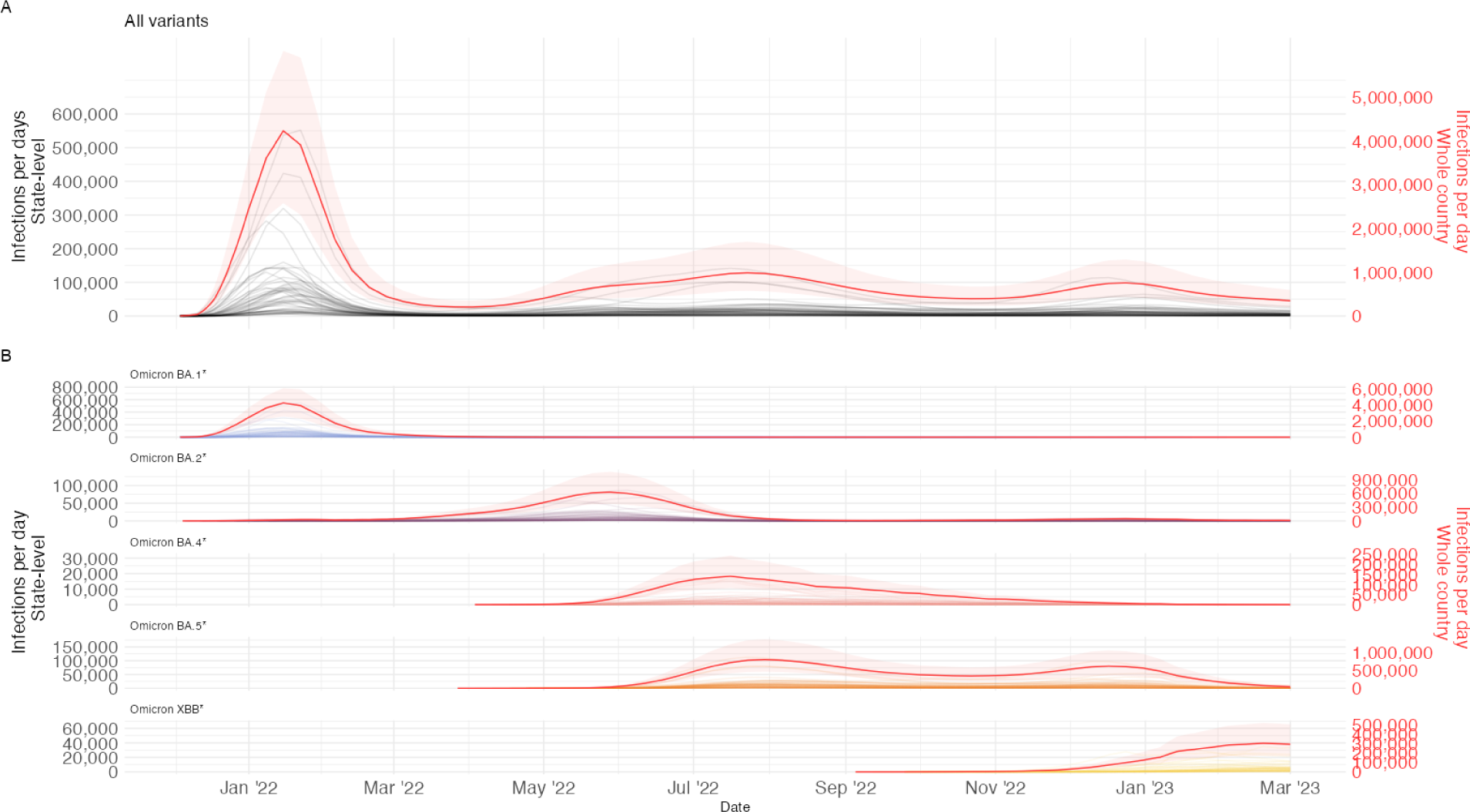
Time series of daily Omicron variant infections across the entire United States. The left y-axis, in black, is the state-level, and the right y-axis, in red, is the national scale. Note that the scale of the y-axis differs between time series for each variant. The red shading is the 95% Credible Interval (CrI) for the national estimate. The state-level 95% CrI for each of the variant infection estimates are provided in **Table S3.** Time series of infection estimates for all variants. The gray lines are infection estimates per state and the red line is the mean infection estimates per day for the whole US. **A)** Time series of infection estimates for each variant. The gray lines are infection estimates per state and the red lines are the mean of infection estimates per day for the whole US. The scales differ by each variant subplot, as each variant had a different size of total infection per day.

Reported cases, hospitalizations, and deaths provide an incomplete picture of the status of the COVID-19 pandemic since the majority of infections are asymptomatic. We address this by using infection estimates from *covidestim* (19), a nowcasting model that generates daily infection estimates while correcting for under-reporting and notification delays (**Fig. 1A**). We then sorted the SARS-CoV-2 sequences for all 50 states and the District of Columbia and binned the lineages into variant categories - BA.1*, BA.2*, BA.4*, BA.5*, and XBB* (**Table S1**). Combining these two sets of analytic outputs, we calculated the daily frequencies of each Omicron variant (**Fig. S1**). We used this information to estimate the number of daily variant-specific infections via a spline interpolation (**Fig. 1B**). For more details see Materials and Methods.

We identified three peaks of infections in 2022 associated with the prevalence of distinct variants, one period in the winter, one in spring to early summer, and one in the late fall (**Fig. 1A**). The first Omicron period (BA.1*, December 2021 - January 2022) caused an estimated 4.2 million (95% credible interval [CrI] = 2.6-6.0 million) infections per day at its peak (about 1.25% of the US population being infected per day) (**Fig. 1B, Tables 1** and **S2**). In total, we estimate that BA.1* caused approximately 169 million infections (95% CrI = 97-249 million) in the US during this wave (**Table 1**). The second Omicron period started in April 2022 (>2% frequency) with the emergence of Omicron BA.2* and lasted until November 2022 (<2% frequency) with the initial emergence of BA.4* and BA.5*. These variant-specific surges peaked at ∼625,000 (BA.2*), ∼140,000 (BA.4*), and ∼800,000 (BA.5*) infections per day in the US. Finally, the third Omicron period, from November 2022 to March 2023, was driven by a resurgence of BA.5* and the emergence of the recombinant variant, XBB*, which peaked at ∼500,000 and ∼300,000 infections per day in the US, respectively.

**Table 1.** Variant-specific Attack Rate, Peak, and Total Infections for the United States.

|  | Variant Categories |  |  |  |  | Overall |
| --- | --- | --- | --- | --- | --- | --- |
|  | Omicron<br>BA.1* | Omicron<br>BA.2* | Omicron<br>BA.4* | Omicron<br>BA.5* | Omicron<br>XBB* |  |
| Attack Rate mean (max, min) |  |  |  |  |  |  |
|  | 47.7%<br>(57.2%,<br>37.9%) | 17.2%<br>(29.6%,<br>6.4%) | 4.4% (6.7%,<br>2.3%) | 35.7%<br>(47.9%,<br>24.3%) | 9.1% (15.6%,<br>3.4%) |  |
| Peak Infections |  |  |  |  |  |  |
| Peak number<br>of daily<br>infections<br>(95% CrI) | 4,204,319,<br>(2,558,883,<br>6,012,892) | 625,656,<br>(366,130,<br>1,065,860) | 140,772,<br>(81,506,<br>243,055) | 799,728,<br>(462,471,<br>1,368,620) | 303,653,<br>(172,254,<br>518,757) |  |
| Total Infections |  |  |  |  |  |  |
| Total<br>infections(95<br>% CrI) | 169,296,672<br>(96,811,952,<br>249,315,168) | 57,105,922<br>(30,272,041,<br>88,933,250) | 15,557,243<br>(8,172,750,<br>24,319,495) | 131,313,161<br>(69,034,994,<br>205,025,830) | 31,476,061<br>(16,254,246,<br>49,538,781) | 404,749,058<br>(220,545,98,<br>617,132,524) |

At the state level, we estimated that the daily BA.1* infections peaked at ∼548,000, ∼422,000, ∼318,000, and ∼281,000, for California, Texas, Florida, and New York, respectively. Similar to our national estimates, at these peaks over 1% of the state population was being infected per day. We summarize the total and peak daily infections for each Omicron variant for all 50 states, the District of Columbia, and the whole country **(Tables 1, S2,** and **S3)**.

### Omicron variant attack rates for each state

We used the daily infections to calculate the percent of the population estimated to have been ever infected during each variant wave (variant-specific attack rates) for each US state (**Figs. 2** and **S4**). During the BA.1* wave, states with the highest attack rates - Kentucky (57.2%), Alabama (56.5%), and Louisiana (56.3%) - were concentrated in the southeast, while we estimate the lowest attack rates from Iowa (38.3%), South Dakota (38.0%), and Idaho (42.1%). The highest and lowest state attack rates for the other Omicron variants were as follows: BA.2* highest in Hawaii (30%), lowest in South Dakota (6%); BA.4* highest in North Carolina (6.8%), lowest in Vermont (2.3%); BA.5* highest in Kentucky (48%), lowest in Vermont (24%); XBB* highest in Rhode Island (15.6%), lowest is Arkansas (3.4%; **Fig. 2B, S4**). While Kentucky often had high attack rates and Vermont and South Dakota generally had lower attack rates, we did not detect consistent geographical patterns for each Omicron variant.

**Fig. 2.**
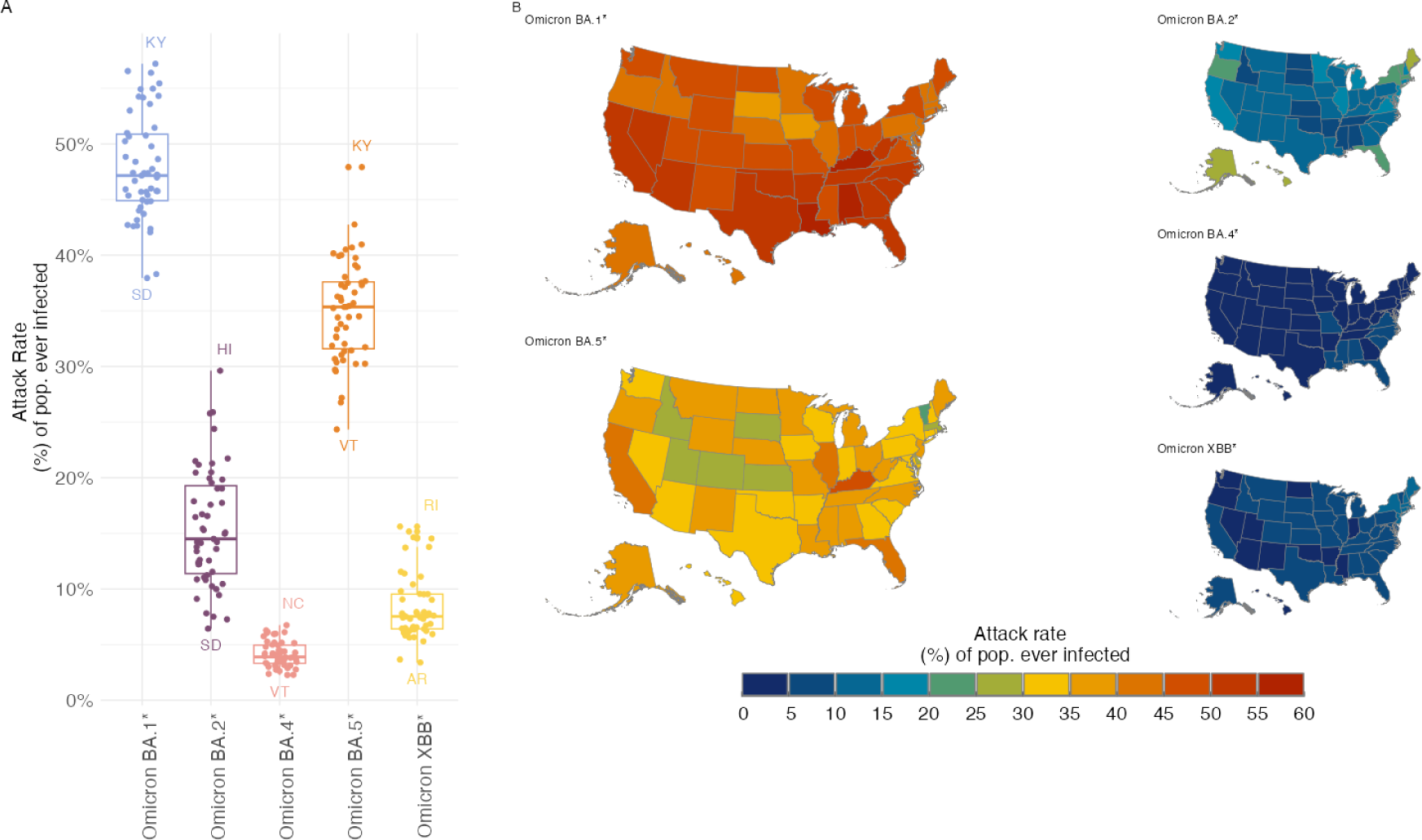
Distribution of attack rate estimates across the United States for each Omicron variant. **A)** Attack rate distribution and state-level attack rate estimates. Each dot is a state attack rate estimate, and the boxplots show the distribution of attack rate values across all states. **B)** Maps of the attack rate estimates. For all the Omicron variants we show the US map, with Alaska and Hawaii placed below. Color on the state map indicates the state-level attack rate value of each variant.

### Variant-specific effective reproduction numbers estimated from across the US

We estimated Omicron variant-specific effective reproductive numbers (R_t_) for each state to gain insight into variant transmission (**Fig. 3)**. We produced variant-specific estimates of R_t_ across all states by applying the *EpiEstim* R package (20,21) functions to our variant-specific daily infection estimates (**Fig. 1B**). For Omicron BA.1*, the median R_t_ across all states started as high as 3 (1.5, 3) (**Table S5**), while the R_t_ estimates for the other variants were smaller. We found similar longitudinal R_t_ estimates for BA.4* and BA.5*, indicating that they were generating similar numbers of secondary cases in the US and thus able to co-exist for several months. This observation suggests that there are variant-specific factors that can impact their relative transmissibility (e.g. immune escape, infectivity), but there are important population factors that also impact infection incidence.

**Fig. 3.**
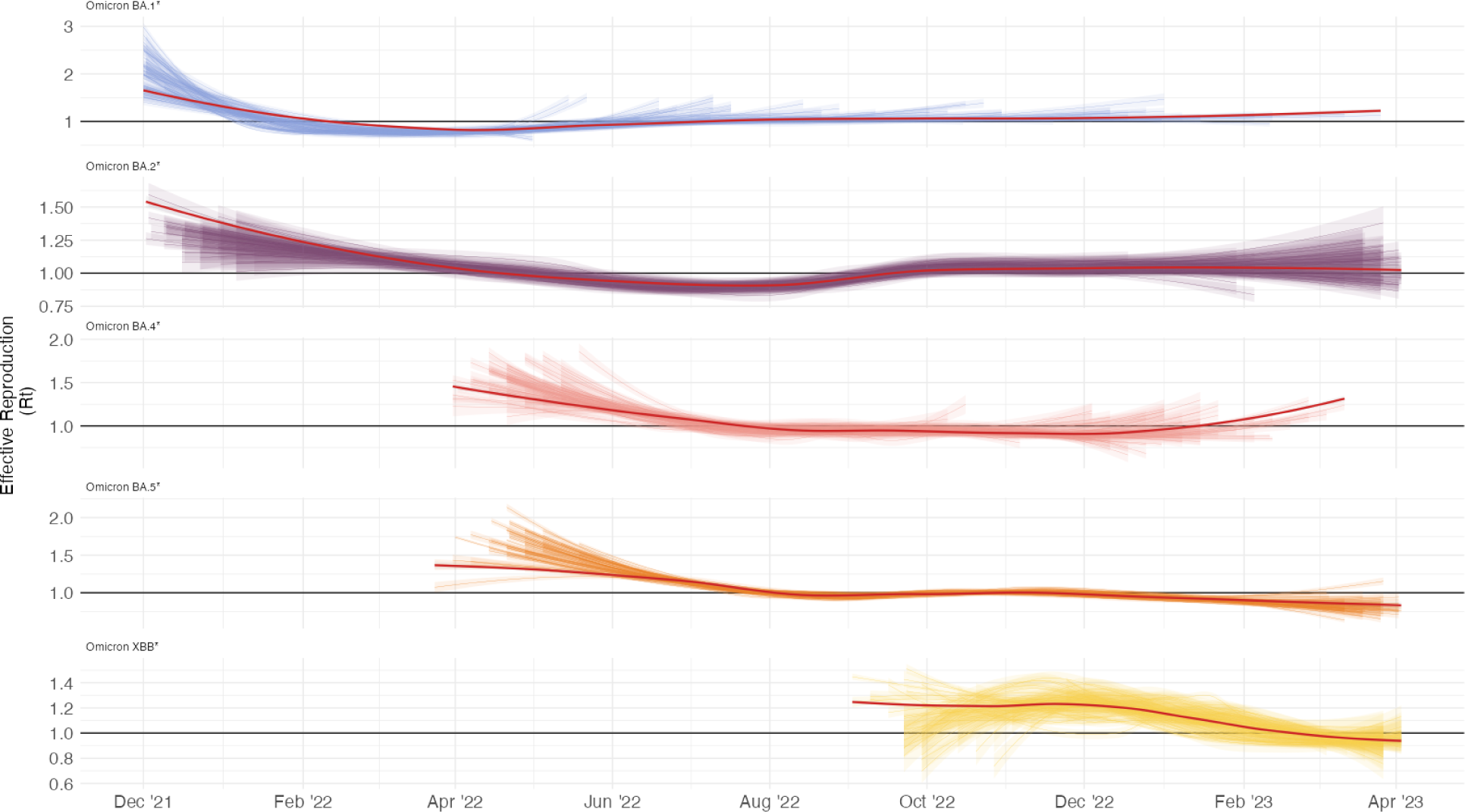
**Time series of variant-specific effective reproductive numbers across all states.** On each facet is depicted the time series for all US states and its confidence interval to the R_t_ estimate. The red line is the national average overall states. To help the visualization we apply over each state R_t_ time series a locally estimated scatterplot smoothing function (LOESS). The y-axes showing the R_t_ values are independently scaled for each variant to highlight changes over time.

### Variant-specific associations between attack rates and social vulnerability

To investigate whether SARS-CoV-2 transmission is associated with population level social vulnerability, we examined correlations between our estimated outcomes and the CDC social vulnerability index (SVI) metric (22). Comparing the state SVI (**Fig. 4A**) to the attack rates for each variant, weighted by the state population sizes (**Fig. 4B**), we found that the Omicron BA.1* (correlation coefficient *R* = 0.56). BA.4* (*R* = 0.3), and BA.5* (*R* = 0.31) attack rates positively correlate with the SVI (**Fig. 4B**). The BA.2* and XBB* emergences occurred immediately following the two largest Omicron waves, BA.1* and BA.5*, respectively. We, therefore, hypothesized that while individuals living in states with higher SVIs have higher exposure rates, they are less susceptible to infection during variant emergence immediately following exposure to a previous novel variant wave.

**Fig. 4.**
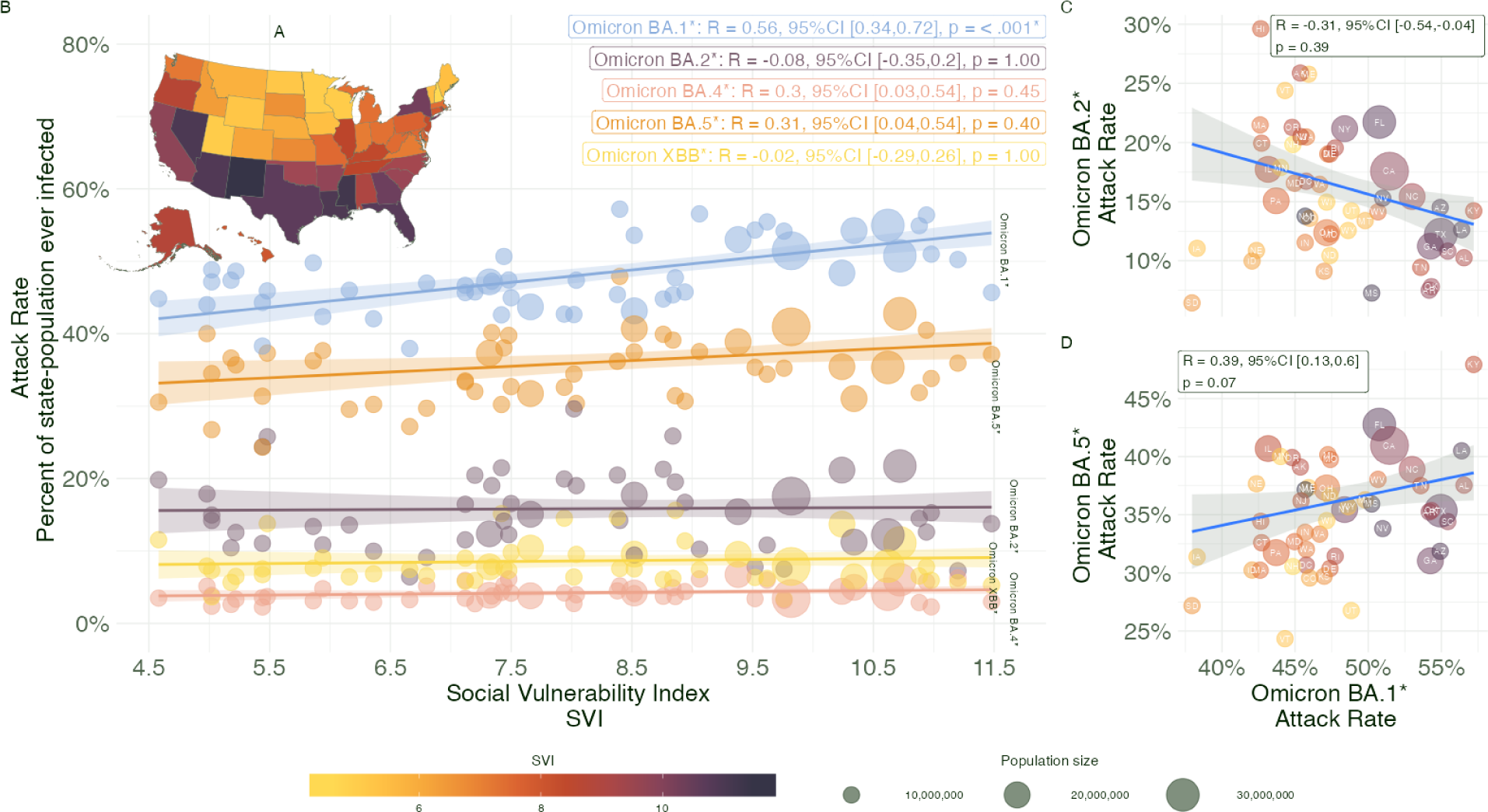
Correlation between variant attack rates and the social vulnerability index. **A)** Map of the SVI for all states, colors correspond to the SVI scores. **B)** Scatterplot between attack rates by variant category and the SVI. Sizes are equivalent to the size of the state population and colors correspond to the variant categories as in the Panel B of Fig. 1. **C)** Scatterplot between the attack rate of Omicron BA.1* and Omicron BA.2*, colors correspond to the SVI quartile, and size is proportional to the state population size. Correlation between the attack rates. **D)** Scatterplot between the attack rate of Omicron BA.1* and Omicron BA.5*, colors correspond to the SVI quartile, and size is proportional to the state population size. Correlation between the attack rates.

To test the hypothesis that states with higher SVI had higher exposure rates, we compared the Omicron BA.1* attack rates to those for BA.2* (peaked ∼4 months after BA.1) and BA.5* (peaked ∼6 months after BA.1). We calculated a negative correlation between the BA.1* and BA.2* attack rates (*R* = −0.31, 95% CI [-0.54, −0.04]; **Fig. 4C**) and a positive correlation between BA.1* and BA.5* (*R* = 0.39, 95% CI [0.13, 0.6]; **Fig. 4D**). States like Kentucky, Louisiana, and Alabama, which are on the higher end of the SVI scale, had attack rates that were relatively low for BA.1*, low for BA.2* attack rates, and high for BA.5*. Four states that did not fit the negative BA.1*-BA.2* correlations were South Dakota, Iowa, Idaho, and Nebraska, all of which had low SVI values and relatively low attack rates for both variants. Thus our analysis supports our hypothesis that variant waves are driven by opposing forces of social vulnerability that govern exposure rates and population susceptibility following previous outbreaks.

## Discussion

We investigated the Omicron variant-specific infection dynamics across all US states, estimating daily infections, attack rates, and effective reproduction numbers. By combining sequencing data with infection estimates, we aimed to disambiguate infection dynamics during periods of strain replacement and when variants were co-circulating, revealing features of the epidemic that could not be inferred from the reported epidemiological data alone.

We found that Omicron variants were responsible for approximately 404 million (95% CrI = 221-617 million) infections across the US from December 2021 to March 2023, including approximately 169 million during the BA.1* wave. The transmission dynamics of variants differed markedly: BA.1* emerged as a genetically distinct (3,23–25) variant which caused large rapid epidemics, especially in states with a higher degree of social vulnerability. Subsequent Omicron variants, while able to both co-circulate and eventually outcompete extant strains, spread at lower levels and often for longer durations, exhibiting much weaker association with social vulnerability measures than the BA.1* variant. These findings reveal the complex interplay between viral evolution, population susceptibility (driven by previous infections and population-level immunity), and social factors that affect the risk of exposure and infection.

The validity of our estimated variant-specific infections, attack rates, and effective reproduction numbers depends on several assumptions (20,21,26,27). The state-level estimates of total infections (i.e. not stratified by variant) were obtained from a published Bayesian nowcasting model which used publicly available time series of COVID-19 case notifications, hospitalizations, and deaths, accounting for effective population immunity. These estimates are calibrated to hospitalization and death data, accounting for delays associated with disease progression and estimates of infection hospitalization and infection fatality ratios (17). The model maintains two sets of assumptions, before and after the introduction of the Omicron variants. The pre-Omicron model does not allow for reinfection or waning of immunity, while the Omicron-era model allows for waning of immunity after infection. Because the underlying mathematical model uses a weekly spline function to model the transmission rates, no explicit assumptions were made about the transmissibility of each variant. Rather, the model allows the transmissibility of circulating variants to vary over time, while the infection-hospitalization ratio remains fixed. By using publically available SARS-CoV-2 sequencing data from GISAID to estimate variant frequencies at the state level, we were able to disaggregate the total number of infections into variant-specific incidence in the current analysis. As such, we assumed the sequencing was done at random within states. We also note that our analysis of the association between state-level attack rates and state-level SVI has the potential for ecological fallacy and should thus be interpreted with caution.

Our findings align with data from blood donors (28) and another modeling study in China (29). The prevalence of anti-spike and anti-nucleocapsid antibodies (infection-induced and hybrid-induced) in the blood donor sample rose from 20.9% in April - June 2021 (Pre-Omicron), to 54.6% in January - March 2022, and then to 70.3% in July - September 2022. The latter two periods align with our estimates of the Omicron BA.1* and BA.5* waves. After the Omicron BA.5* wave, we estimate a cumulative attack rate of 83.4% of the US population. The China study estimates after the BA.5* introduction in a naive population, that 97% of the population had been infected. The overall attack rate we estimate is larger than the US population, which is explained by reinfections over the Omicron variant waves.

Our findings provide evidence that the dynamic evolution of SARS-CoV-2 variants is a result of the interplay between exposure and immunity to the virus (3,18,30,31). The pandemic’s history has been marked by the initial emergence of highly transmissible variants (3,7,27,32) and the Omicron era is marked with immune escape characteristics (3,30,33), necessitating ongoing adaptations in public health responses. By quantifying infection rates, attack rates, and effective reproduction numbers for different variants across all states, we provide valuable insights that can guide preparedness and resource allocation.

## Materials and Methods

First, we describe the processing of lineage information and how the lineages were summarized into categories. Second, we describe how the variant-specific infection estimates are produced by joining the infection estimate time series and variant frequency time series. Third, we describe the use of a modified version of *EpiEstim* tools to estimate the R_t_ for each of the variants. Lastly, we describe the joint analysis of the attack rate estimates and social vulnerability index scores.

### Data Sources

The GISAID database contains more than 16 million genomes, of which approximately one-third come from US genomic surveillance efforts (11). We processed the metadata and generated counts and frequencies of each variant lineage. Frequencies of variants have been used as a surveillance tool by the Centers for Disease Control and Prevention (CDC) and can give information on new invading variants. The GISAID metadata contains the Pango lineage nomenclature system classification of the genome. We can further distribute lineage information into variant categories by aggregating the major parental lineages and their sublineages into the same category. We categorized those lineages into major lineages categories (which we refer to as “Omicron variants”), such as Omicron BA.1* to incorporate Omicron BA.1 and its sublineages, Omicron BA.2* to Omicron BA.2 and its sublineages, and so on (**Table S1**).

We used the published *covidestim* model data to render weekly variant-specific estimates of infection from December 1, 2021 until May 1, 2023. This model back-calculates infections from the observed case, death, vaccination and hospitalization reports, using assumptions on reporting and progression delays, and a variable probability of case reporting over time. From the beginning of the pandemic until December 1, 2021, reinfections were not assumed to occur and the model was based on case and death reports. After December 1, 2021, infections are back-calculated from case and hospitalization reports, to accommodate the reduced mortality rate under Omicron variants. Furthermore, vaccination reports and assumed waning of immunity are included in the assumptions, and the serial interval and infection mortality rate assumptions are adjusted to match the new disease dynamics (17). The model output is a median of the infection estimates and its 95% credible interval (CrI).

### Lineages collapsing into major lineage categories

We pre-processed the metadata downloaded from GISAID and categorized the Pango lineages into 8 major categories: ‘Omicron BA.1*’, ‘Omicron BA.2*’, ‘Omicron BA.3*’, ‘Omicron BA.4*’, ‘Omicron BA.5*’, ‘Omicron XBB*’, ‘Other Recombinant’ and ‘Other’, see **Table 1** for details on each lineage and its sublineage alias. As for the categories such as ‘Omicron BA.3*’, ‘Other Recombinant’, and ‘Other’, had less than 2% in frequency and we suppressed them from the main analysis. We collapsed all the sub-lineages into major categories, the following table summarizes our categorization. From the categorization, we count and calculate the frequency of each of these categories in every state and week. See **Fig. S1** in the supplementary material for the counts of genomic sequences for the whole US during the studied period, Dec 2021 to May 2023, into the 8 previously mentioned categories.

### Variant-specific estimates by joining genomic frequencies and infection estimate

We summarized the genomic sequence data to align with the *Covidestim* weekly infection estimates. From weekly counts, we calculate the frequency of each of the major variant categories described above. This process guarantees compatibility between the dates of metadata and infection estimates. We filter out frequencies below 2% on a week. By multiplying the frequencies of each category at each state by the number of total infections estimated for each state weekly, we produce estimates of the infections per variant in each state per week. We round the number of infections estimated to an integer number of infections.

In a formula, we have:

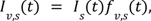

Where the infection estimate time series for the variant *v* at state *s, I_v,s_*(*t*), is given by the infection estimate time series of total infection at state *s*, *Is*(*t*), times the frequencies time series of each variant *s* within the state *s*, *f_v,s_* (*t*). With *f_v,s_* (*t*) > 0. 02 every week.

We interpolate the weekly time series using a b-spline function to produce a daily time series of infections. We repeated the same procedure to the 2.5th and 97.5th quantiles of the infection estimates generated by *Covidestim*. We report the 2.5th and 97.5th quantiles of the posterior distribution trajectories as the lower and upper bound, respectively, for the 95% credible interval (CrI) of the infection estimates. To compare the incidence estimates by each variant, we calculated the cumulative incidence over the epidemic of each variant for all states. The incidence is given as the percent of the population ever infected with the variant in the state.

### Effective reproduction number estimates

The daily time series of each variant in each state was then given to the ‘estimate_R()’ function from the R package *EpiEstim*. To avoid non-converging problems with the model employed by *EpiEstim*, we only parse time series with more than ten days of continuous infection estimates. The model is parametrized using an uncertain Serial Interval (SI) setting, estimating the serial intervals of SARS-CoV-2 (Omicron variant specific) by drawing from two (truncated) normal distributions for the mean and standard deviation of the SI. The truncated normal distribution of the SI is then parametrized with a mean of 3.5 (1–6 days).

### Variant-specific attack rate

From the variant-specific infection estimates we can calculate the variant-specific attack rate (AR), *i.e*. the proportion of state-population ever infected on each variant *v* wave. We calculate the AR by summing all the infections for a specific variant *v* at state *s* over the period of the study:

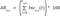

Where *Inc_v,s_* (*t*) is the incidence of variant *v* at state *s*, given by:

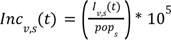

### R_t_ ratios per variant for each state

We calculated the R_t_ ratio for pairs of variants to compare the R_t_ values between each variant across states (**Fig. S6**). We created pairs based on temporal succession; for time points with two or more variants co-circulating, we divided the succeeding variant time series by the preceding variant time series (**Fig. S6A**). In all pairs of succession, we found the average R_t_ ratio was greater than 1 as expected, consistent with the observation that succeeding variants were capable of invasion (**Fig. S6B** and **Fig. S5**).

We estimate the advantage of one variant over another by taking the R_t_ ratios during their period of coexistence. From the R_t_ ratios, we can classify two different periods to the succession of Omicron variants. Periods of complete clearance of the previous variants are marked with higher R_t_ ratios, as for the R_t_ between BA.2*/BA.1* and XBB/BA.5* (**Fig. 3**.). Conversely, we see periods of coexistence of more than one variant have smaller R_t_ ratios, e.g., the ratios between BA.4*/BA.2*, BA.5*/BA.2* and BA.5*/BA.4*. The median R_t_ values of BA.2*, across the US, were almost 20% higher than the R_t_ values of BA.1*, and to XBB* distribution of R_t_ values it was more than 20% bigger than the R_t_ values of BA.5*. In summary, variants with comparable higher R_t_ (BA.2* and XBB*) values to their predecessor, can completely invade the dominant variants. See **Fig. S6.** for the R_t_ ratios for all states.

### Assessing the association between state-wide Social Vulnerability Index and variant-specific AR

The social vulnerability index (SVI) is a metric compiled by the CDC summarizing the social conditions that may affect the outcome in the face of disasters, such as infectious disease outbreaks (22) (**Fig. 4**). The SVI is a summary metric, incorporating 4 main domains: socioeconomic status; household characteristics; racial and ethnic minority status; and housing type and transportation. States that are high on the SVI scale tend to have larger populations and are primarily concentrated in the southern half of the US (**Fig. 4A**). Originally the index was compiled at the census tract and county level; we have aggregated them by state to be able to use it with the state-level estimates of infections by variant.

We calculated the correlation between the SVI and the state-level variant-specific AR using Pearson correlation. For each of the variant-specific correlations between the SVI and the AR, we calculate statistical significance with Bonferroni correction for multiple testing.

## Data availability

The findings of this study are based on metadata associated with 3,103,250 sequences available on GISAID from September 1st, 2021 up to April 22, 2023, and accessible at https://doi.org/10.55876/gis8.231023hd (GISAID Identifier: EPI_SET_231023hd). All genome sequences and associated metadata in this dataset are published in GISAID’s EpiCoV database. To view the contributors of each sequence with details such as accession number, Virus name, Collection date, Originating Lab and Submitting Lab, and the list of Authors, visit https://doi.org/10.55876/gis8.231023hd

The *covidestim* model uses publicly available data on case and death reports from Johns Hopkins University and the CDC, vaccination data from the CDC and hospitalization data from Healthdata.gov (34–36). The script to join these data sources is available on Github (https://www.github.com/covidestim/covidestim-sources), and the full description on how the data is modeled is available in the linked publications (16,17,19).

Both the input data to the model, and the produced estimates used for this analysis are available on Github (https://www.github.com/covidestim/data-archive).

## Code availability

The pipeline used to calculate the variant-specific infections, attack rates, R_t_, R_t_ ratio, and SVI comparison is available on the following GitHub repository: https://github.com/rafalopespx/Variant_infections_rate

## Data Availability

All data produced are available online at: https://github.com/rafalopespx/Variant_infections_rate

## Acknowledgments

We gratefully acknowledge T. Thornhill for the discussion and helpful insights with the Social Vulnerability Index and P. Jack and S. Taylor for technical support, and the authors from the originating laboratories responsible for obtaining the specimens, as well as the submitting laboratories where the genomic data were generated and shared via GISAID, on which this research is based. To view the contributors of each sequence with details such as accession number, Virus name, Collection date, Originating Lab and Submitting Lab, and the list of Authors, visit https://doi.org/10.55876/gis8.231023hd. All plots use color palettes from the ‘MetBrewer’ R package https://github.com/BlakeRMills/MetBrewer. This project is supported by Cooperative Agreement NU38OT000297 from the Centers for Disease Control and Prevention (CDC) and the Council of State and Territorial Epidemiologists (CSTE), SHEPheRD Contract 200-2016-91779 from the CDC, and the CDC Broad Agency Announcement Contract 75D30122C14697. This work does not necessarily represent the views of the CDC or CSTE.

## Author contribution

Conceptualization: RL, TC, NDG

Methodology: RL, KP, FK, TC, NDG

Investigation: RL, SR, AH

Visualization: RL

Funding acquisition: TC, NDG

Supervision: JAS, NAM, TC, NDG

Writing – original draft: RL, TC, NDG

Writing – review & editing: all authors

## Delration of Interests

NDG is a paid consultant for BioNTech.

---

**Fig. S1.**
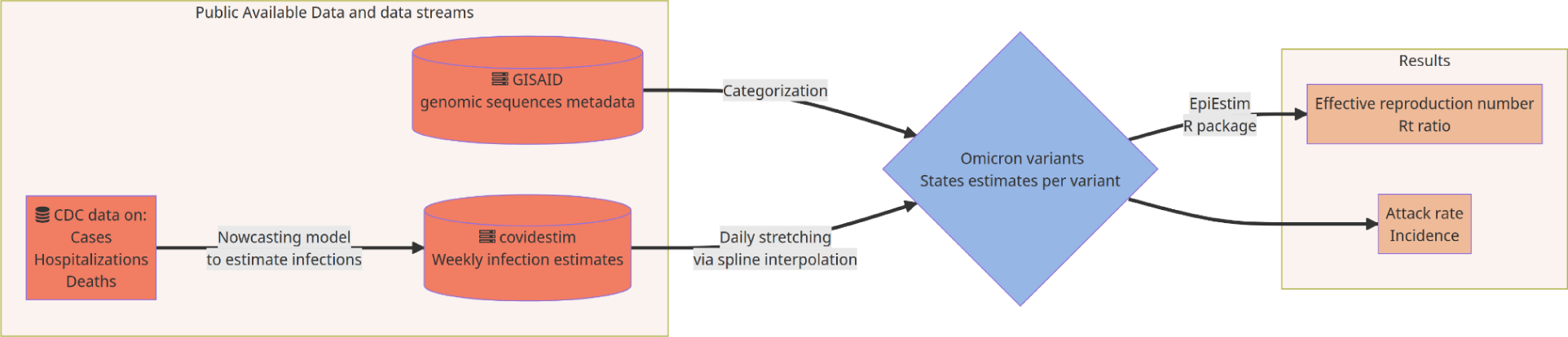
Flowchart of the process of joining genomic and epidemiological data streams. From the genomic sequences metadata GISAID and infection estimates from *covidestim*, we produced infection estimates by multiplying the frequencies of each Omicron variant by the infection estimates. Those estimates are then imputed to *EpiEstim* functions to produce variant-specific effective reproduction numbers, R_t,_ and state attack rates.

**Fig. S2.**
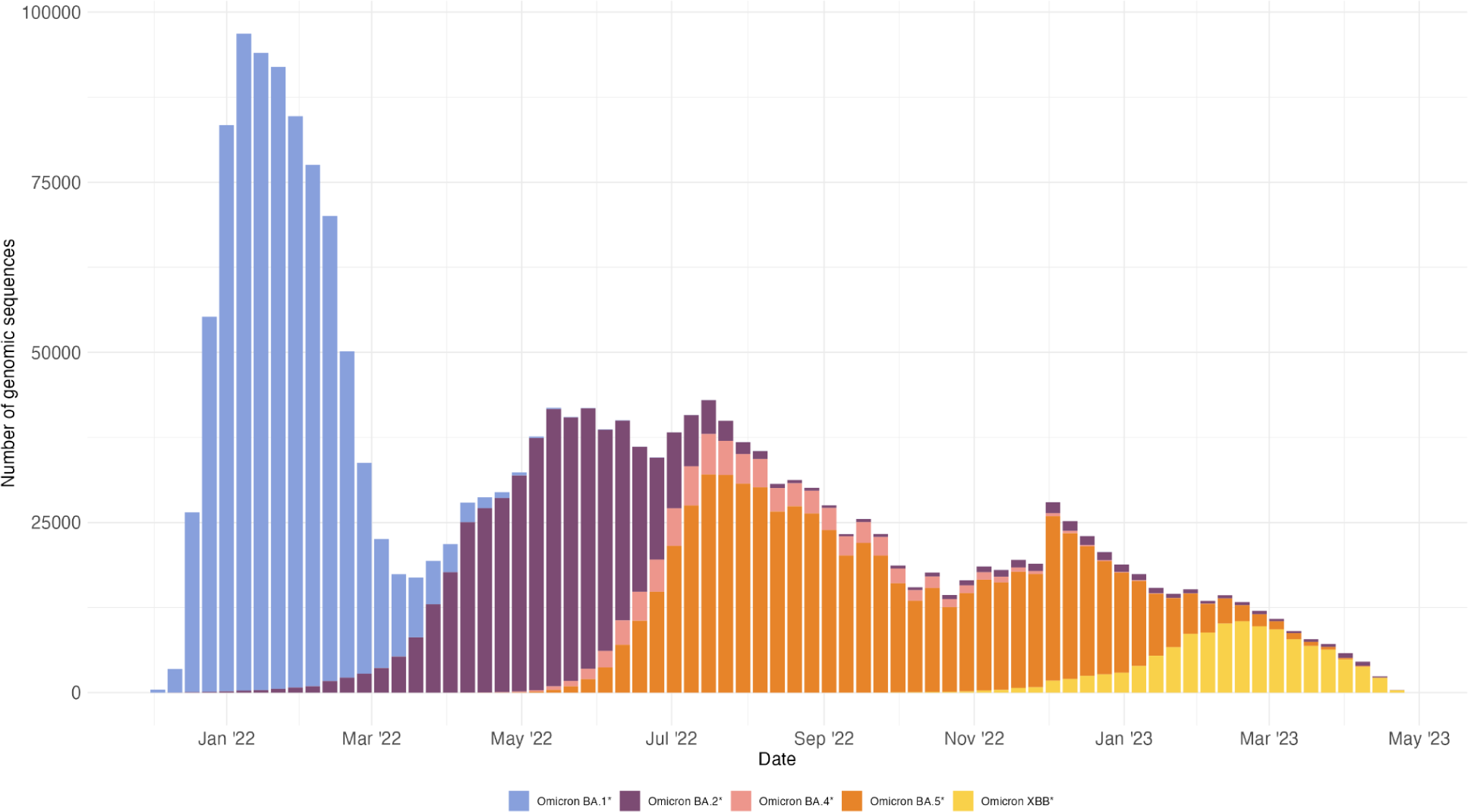
Number of genomic sequences per variant category per week during the period of December 1st, 2021 to May 1st, 2023, to the whole country. From the GISAID metadata, we calculate the amount of sequences deposited to the database per week, during the analyzed period. Each bar is a week of the period and the filling of the bar is the frequency of each variant during that week. It is possible to see the pattern of succession of variants over the year.

**Fig. S3.**
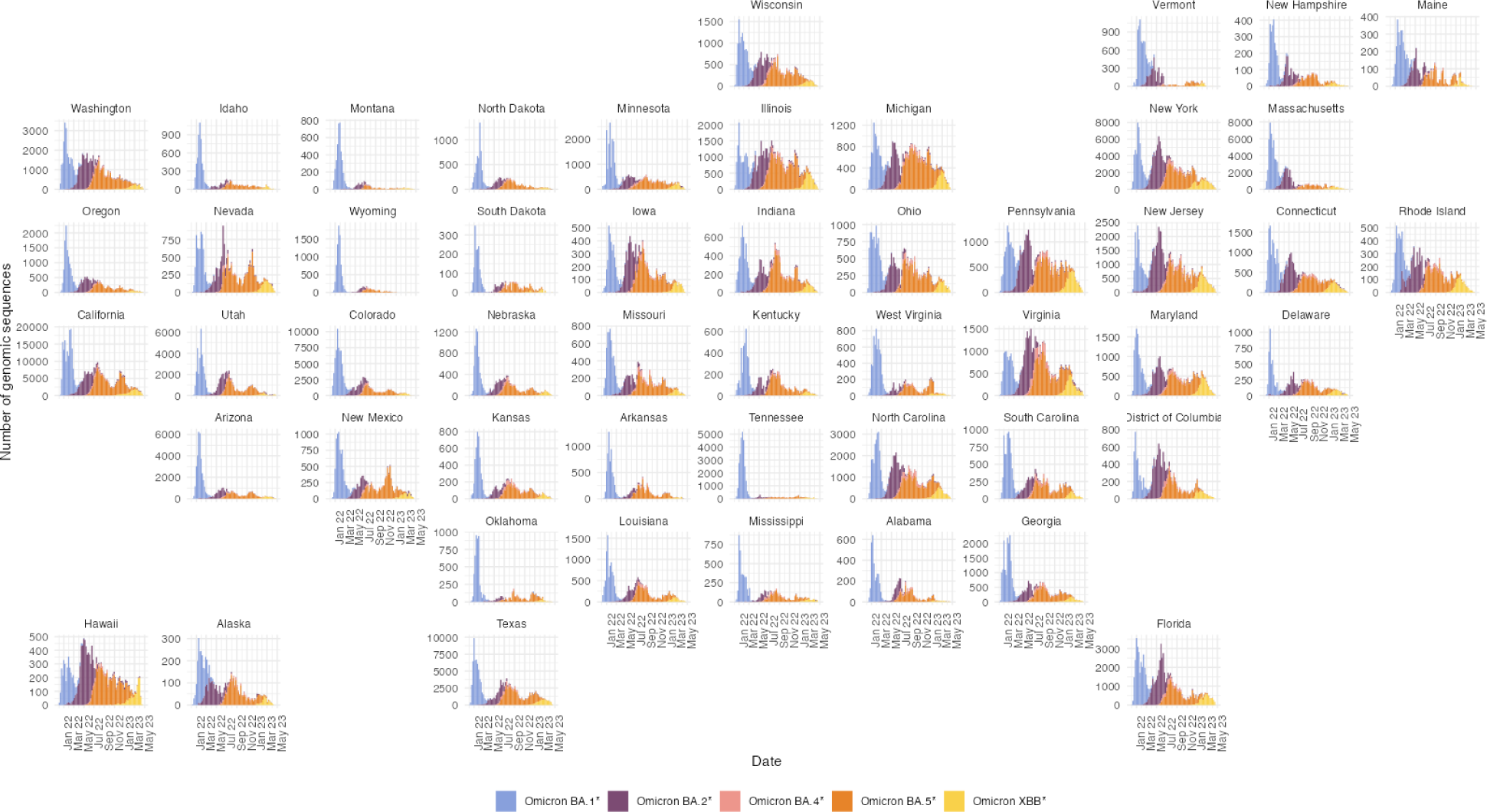
Number of genomic sequences per variant category per week during the period of December 1st, 2021 to May 1st, 2023, to all individual states. From the GISAID metadata, we calculate the amount of sequences deposited to the database per week per state, during the analyzed period. Each bar is a week of the period and the filling of the bar is the frequency of each variant during that week. It is possible to see the pattern of succession of variants over the year.

**Fig. S4.**
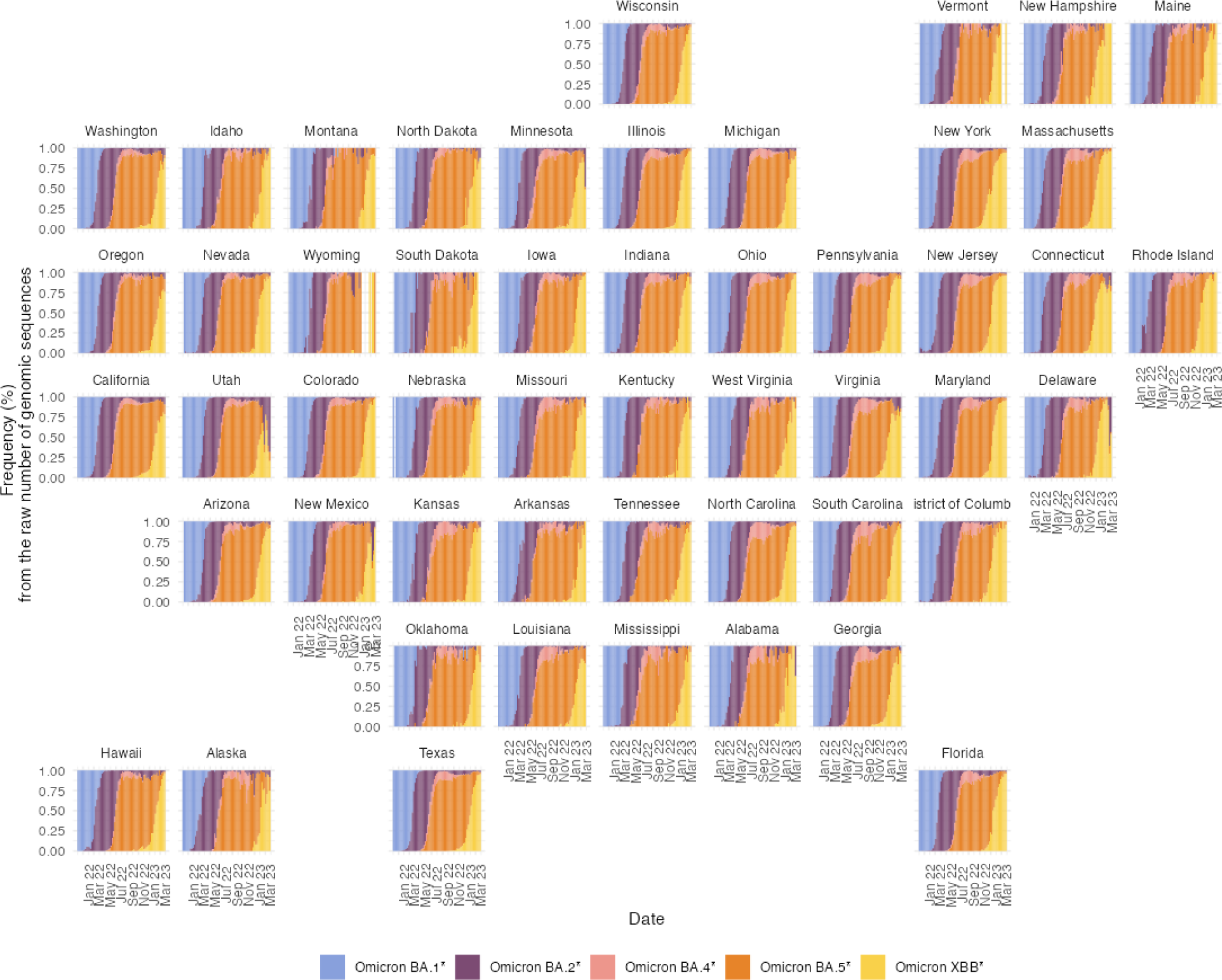
Frequency from the raw number of genomic sequences per variant category, during the period ranging from December 1st, 2021 to May 1st, 2023, over all the individual states. From the GISAID metadata, we calculate the amount of sequences deposited to the database per week, during the analyzed period. Each bar is a week of the period and the filling of the bar is the frequency of each variant during that week. It is possible to see the pattern of succession of variants over the year.

**Fig. S5.**
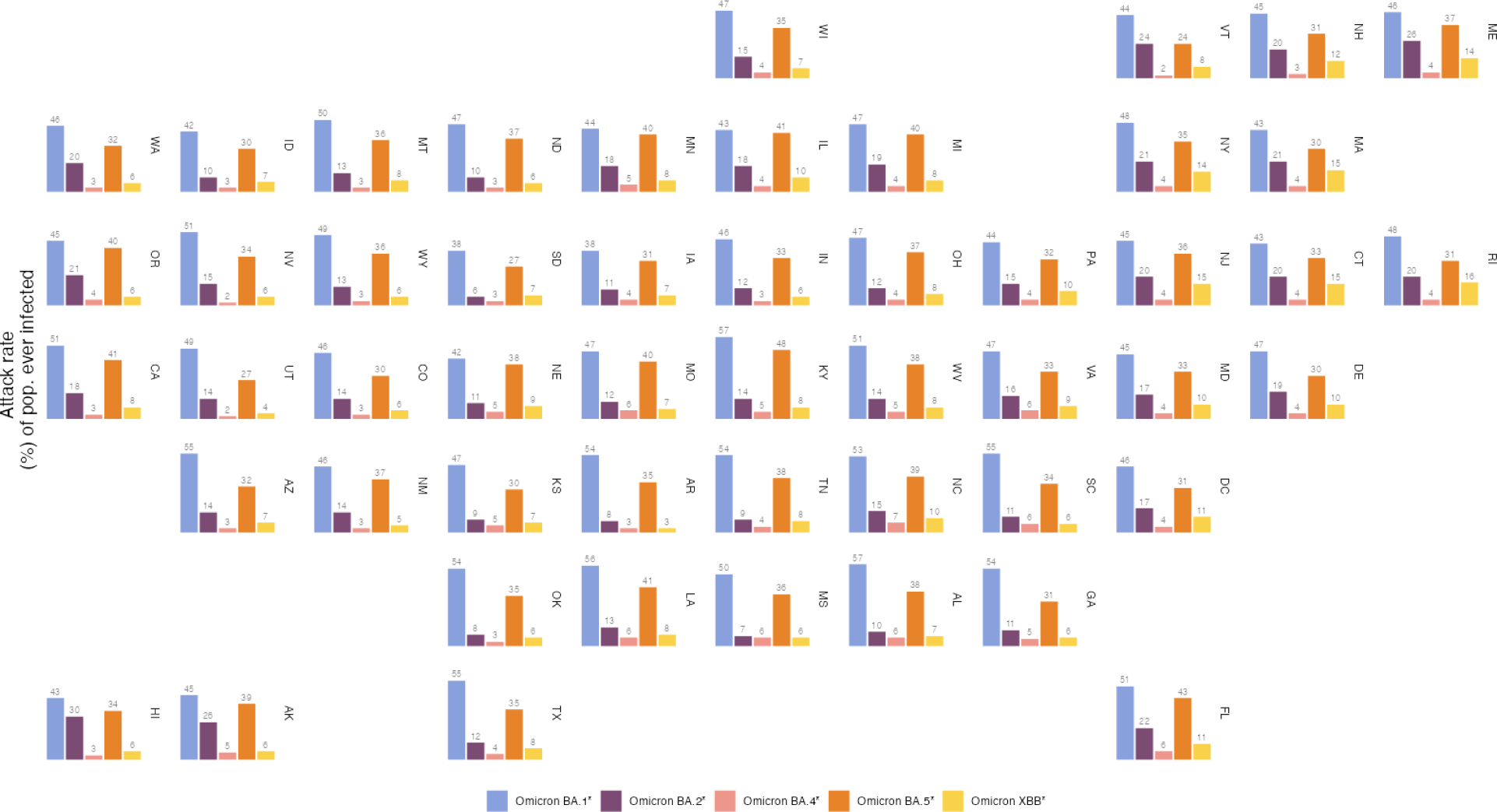
Attack rate per each variant category for all individual states. Bar chart to the variant-specific attack rates estimates in the layout of the US states. Each chart is the attack rates of the variants with the corresponding color. The double-letter state abbreviation is displayed on the right side of each subchart.

**Fig. S6.**
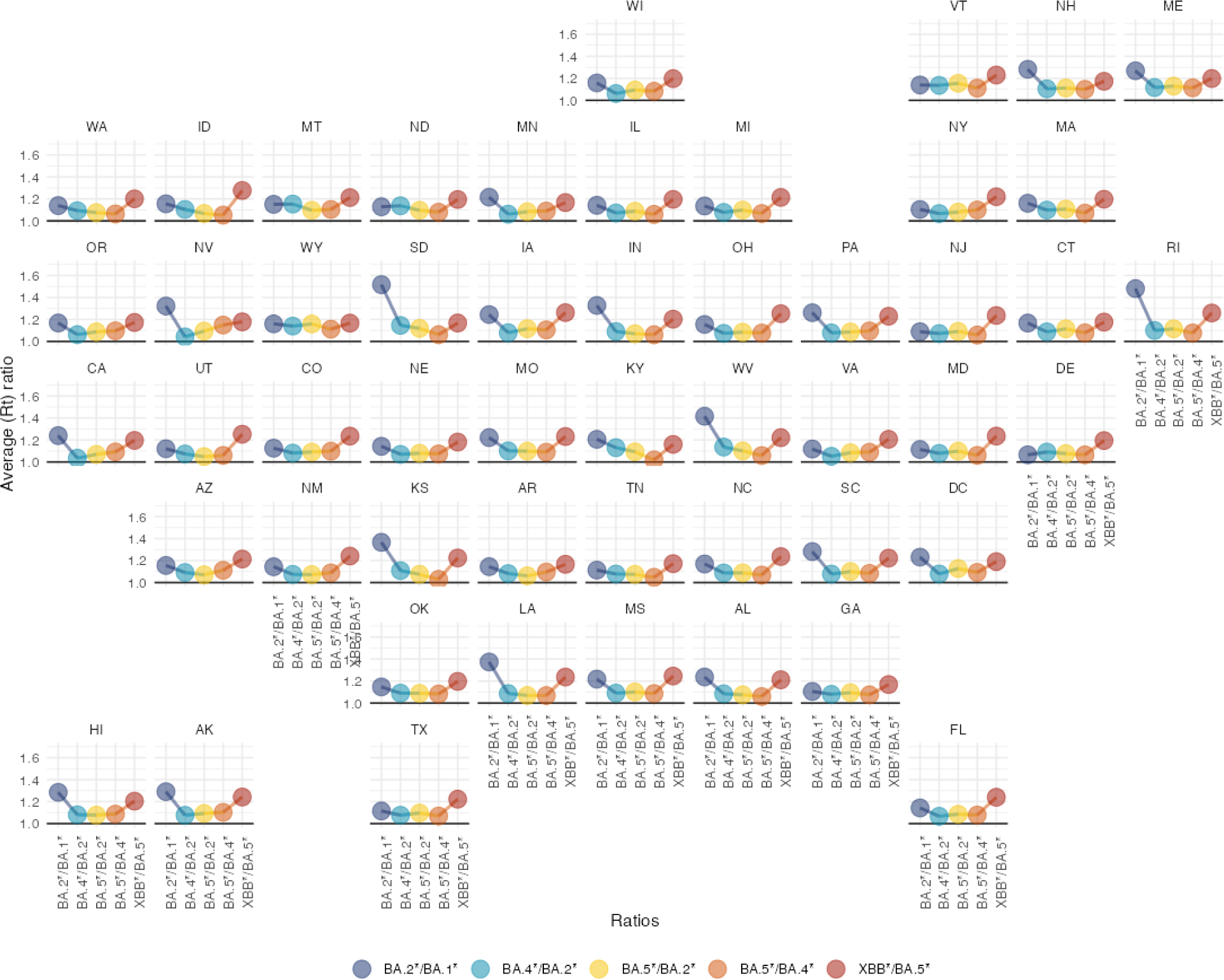
Effective reproduction number (R_t_) ratios to each pair of succeeding variants by each state overall. The R_t_ ratio is calculated by dividing the average R_t_ of the predecessor variant by the successor variant. When the slope rises it means the entering variant has a larger value of average R_t_ over the predecessor variant.

**Fig. S7.**
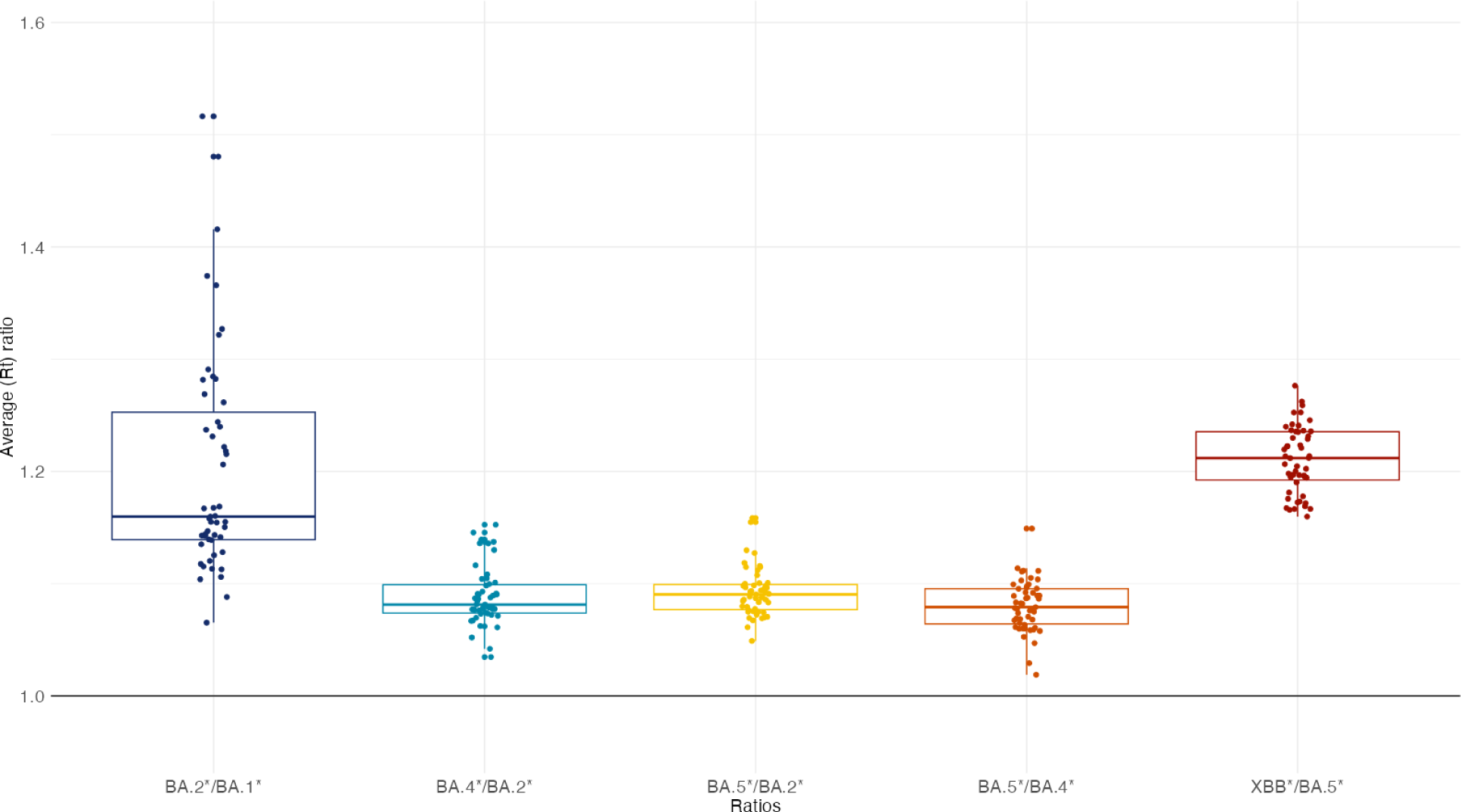
The ratio between variant-specific R_t_ boxplot and dots to the state-specific ratios. Dots are the state-level R_t_ ratio and the boxplot is the distribution over all the states. The pairs of variants are chosen as the succeeding history of variants throughout 2022. After the BA.1* wave and before the XBB* Introduction to the US, the R_t_ ratios are pretty similar, which was a period of coexistence of variants. The BA.2*/BA.1* and BA.5*/XBB* are significantly higher, and mark the complete clearance of the previous variant, respectively BA.1* and XBB*.

**Table S1.** Categorization of Pango lineages and sublineages alias.

| Category | ‘Omicron BA.1*’ | ‘Omicron BA.2*’ | ‘Omicron BA.3*’ | ‘Omicron BA.4*’ | ‘Omicron BA.5*’ | ‘Omicron XBB*’ | ‘Recombinant’ | ‘Other’ |
| --- | --- | --- | --- | --- | --- | --- | --- | --- |
| Pango lineage | BA.1 or B.1.1.529.1 | BA.2 or B.1.1.529.2 | BA.3 or B.1.1.529.3 | BA.4 or B.1.1.529.4 | BA.5 or B.1.1.529.5 | XBB | X, excluding XBB | Any other |
| Sublineages alias | BD.1 | B["G" "H" "J" "L" "M" "N" "R" "S" "Y"] or | No alias for sublineages | C["S"] or D["C"] | B["E" "F" "K" "Q" "T" "U" "V" "W" "Z"] or | No alias for sublineages | No alias for sublineages | No alias for sublineages |
|  |  | C["A" "B"<br>"H" "J"<br>"M" "V"]<br>or<br>D["D" "S"<br>"V"] or<br>E["J" "P"]<br>or F["J"] |  |  | C["C"<br>"D" "E"<br>"F" "G"<br>"K" "L"<br>"N" "P"<br>"Q" "R"<br>"T" "U"<br>"W" "Y"<br>"Z"] or<br>D["A"<br>"B" "E"<br>"F" "G"<br>"H" "J"<br>"K" "L"<br>"M" "N"<br>"P" "Q"<br>"R" "T"<br>"U" "W"<br>"Y" "Z"]<br>or<br>E["A"<br>"C" "D"<br>"E" "F"<br>"H" "N"<br>"Q" "R"<br>"S" "T"<br>"V" "W"<br>"Y" "Z"]<br>or<br>F["A" "B"<br>"C" "F"] |  |  |  |

**Table S2.** Peak of infections by variant categories.

|  | Variant Categories |  |  |  |  |
| --- | --- | --- | --- | --- | --- |
|  | Omicron BA.1* | Omicron BA.2* | Omicron BA.4* | Omicron BA.5* | Omicron XBB* |
| USA |  |  |  |  |  |
| **Peak (95% Crl)** | 4,204,319,<br>(2,558,883,<br>6,012,892) | 625,656,<br>(366,130,<br>1,065,860) | 140,772,<br>(81,506,<br>243,055) | 799,728,<br>(462,471,<br>1,368,620) | 303,653,<br>(172,254,<br>518,757) |
| State |  |  |  |  |  |
| **Alabama** | 80,182 (47,369,<br>112,479) | 6,723 (3,580,<br>10,492) | 4,875 (2,530,<br>7,635) | 16,326 (8,527,<br>25,631) | 6,530 (3,381,<br>10,225) |
| **Alaska** | 9,613 (5,302,<br>14,510) | 2,068 (1,117,<br>3,182) | 465 (253, 713) | 2,942 (1,595,<br>4,514) | 858 (455,<br>1,330) |
| **Arizona** | 103,856<br>(63,738,<br>143,861) | 14,316 (7,904,<br>21,918) | 2,715 (1,494,<br>4,127) | 21,139 (11,237,<br>32,554) | 6,405 (3,408,<br>9,832) |
| **Arkansas** | 50,336 (31,764,<br>69,471) | 2,400 (1,369,<br>3,573) | 1,186 (681,<br>1,770) | 8,830 (5,071,<br>13,182) | 1,831 (1,076,<br>2,703) |
| **California** | 557,489<br>(341,862,<br>782,556) | 95,148 (52,830,<br>145,546) | 18,138 (10,020,<br>27,667) | 125,358<br>(69,079,<br>193,731) | 37,647 (20,103,<br>57,546) |
| **Colorado** | 76,557 (48,741,<br>105,504) | 12,656 (7,609,<br>18,493) | 2,259 (1,308,<br>3,341) | 13,696 (7,733,<br>20,466) | 5,427 (3,099,<br>8,177) |
| **Connecticut** | 43,946 (27,318,<br>60,374) | 11,349 (6,507,<br>16,918) | 1,516 (863,<br>2,265) | 6,990 (3,850,<br>10,572) | 7,253 (4,116,<br>10,801) |
| **Delaware** | 13,451 (8,461,<br>18,504) | 3,039 (1,792,<br>4,442) | 438 (255, 646) | 2,243 (1,313,<br>3,280) | 1,265 (741,<br>1,843) |
| **District of Columbia** | 11,091 (6,650,<br>15,720) | 1,720 (890,<br>2,753) | 355 (183, 560) | 1,682 (902,<br>2,633) | 1,199 (606,<br>1,926) |
| **Florida** | 321,120<br>(180,568,<br>464,205) | 80,828 (34,453,<br>140,930) | 20,522 (8,922,<br>35,349) | 93,418 (39,452,<br>161,334) | 31,583 (13,684,<br>54,676) |
| **Georgia** | 159,773<br>(89,313,<br>230,749) | 16,147 (8,423,<br>25,120) | 6,634 (3,272,<br>10,511) | 31,122 (15,669,<br>49,259) | 9,163 (4,558,<br>14,450) |
| **Hawaii** | 19,096 (12,542,<br>26,064) | 6,743 (4,053,<br>9,836) | 525 (318, 758) | 3,921 (2,392,<br>5,658) | 1,721 (963,<br>2,601) |
| **Idaho** | 17,949 (10,225,<br>26,484) | 2,507 (1,379,<br>3,798) | 1,014 (564,<br>1,525) | 4,328 (2,361,<br>6,496) | 1,997 (1,053,<br>3,064) |
| **Illinois** | 146,721<br>(86,668,<br>212,686) | 33,088 (16,835,<br>53,212) | 5,987 (3,059,<br>9,572) | 35,119 (17,565,<br>56,728) | 16,852 (8,469,<br>27,173) |
| **Indiana** | 84,919 (48,758,<br>124,303) | 9,738 (5,462,<br>14,721) | 2,672 (1,482,<br>4,044) | 18,132 (10,096,<br>27,476) | 5,728 (3,213,<br>8,654) |
| **Iowa** | 32,368 (18,300,<br>48,088) | 4,887 (2,620,<br>7,553) | 1,468 (796,<br>2,252) | 7,617 (4,105,<br>11,718) | 4,089 (2,149,<br>6,364) |
|  | Omicron BA.1* | Omicron BA.2* | Omicron BA.4* | Omicron BA.5* | Omicron XBB* |
| **Kansas** | 38,318 (22,887, 54,483) | 3,556 (2,064, 5,272) | 2,150 (1,251, 3,193) | 6,125 (3,522, 9,115) | 3,056 (1,696, 4,627) |
| **Kentucky** | 58,941 (35,903, 82,128) | 9,077 (5,092, 13,689) | 2,421 (1,345, 3,635) | 18,105 (9,887, 27,601) | 4,984 (2,780, 7,498) |
| **Louisiana** | 76,514 (48,412, 106,023) | 9,231 (5,308, 13,842) | 3,466 (1,935, 5,290) | 15,276 (8,585, 23,280) | 5,480 (3,074, 8,324) |
| **Maine** | 10,704 (5,563, 16,480) | 6,207 (3,173, 9,679) | 561 (293, 865) | 3,648 (1,894, 5,573) | 3,295 (1,702, 5,118) |
| **Maryland** | 83,356 (53,404, 113,293) | 16,156 (9,322, 24,307) | 2,484 (1,375, 3,819) | 13,879 (7,680, 21,416) | 8,350 (4,498, 13,044) |
| **Massachusetts** | 86,114 (54,662, 120,852) | 24,465 (14,110, 36,772) | 2,681 (1,450, 4,137) | 13,242 (7,307, 20,031) | 12,965 (6,938, 20,023) |
| **Michigan** | 117,269 (73,085, 164,533) | 30,366 (16,847, 46,137) | 3,041 (1,673, 4,658) | 25,556 (13,749, 39,398) | 12,241 (6,687, 18,714) |
| **Minnesota** | 63,291 (36,011, 91,500) | 15,655 (8,310, 24,233) | 3,226 (1,756, 4,934) | 13,960 (7,403, 21,740) | 7,029 (3,604, 11,194) |
| **Mississippi** | 44,990 (26,330, 64,764) | 3,129 (1,585, 5,019) | 2,975 (1,553, 4,674) | 9,379 (4,820, 15,136) | 2,236 (1,120, 3,606) |
| **Missouri** | 80,372 (44,259, 118,202) | 10,223 (5,393, 15,688) | 4,489 (2,371, 6,929) | 15,092 (8,016, 23,262) | 6,564 (3,359, 10,232) |
| **Montana** | 12,415 (7,200, 17,745) | 2,320 (1,286, 3,478) | 893 (490, 1,339) | 2,594 (1,435, 3,897) | 1,392 (748, 2,113) |
| **Nebraska** | 20,631 (10,612, 31,697) | 3,127 (1,405, 5,288) | 1,248 (591, 2,066) | 6,412 (3,003, 10,728) | 3,621 (1,673, 5,939) |
| **Nevada** | 46,695 (26,554, 67,458) | 7,282 (3,789, 11,436) | 1,347 (670, 2,179) | 11,321 (5,521, 18,289) | 2,449 (1,270, 3,818) |
| **New Hampshire** | 15,072 (8,165, 22,673) | 4,573 (2,122, 7,523) | 658 (323, 1,050) | 3,084 (1,459, 4,987) | 1,910 (916, 3,086) |
| **New Jersey** | 129,431 (84,798, 175,301) | 25,503 (14,722, 38,458) | 4,113 (2,417, 6,097) | 20,953 (11,991, 31,485) | 20,233 (11,420, 30,806) |
| **New Mexico** | 26,044 (16,304, 36,133) | 4,557 (2,648, 6,670) | 899 (527, 1,311) | 5,621 (3,375, 8,163) | 2,151 (1,217, 3,251) |
| **New York** | 279,829 (188,920, 369,299) | 62,000 (35,089, 94,159) | 8,599 (4,869, 13,046) | 43,532 (24,380, 66,294) | 33,017 (18,181, 50,426) |
| **North Carolina** | 145,495 (82,058, 210,783) | 23,801 (12,104, 37,938) | 7,066 (3,474, 11,378) | 31,189 (15,348, 50,306) | 13,495 (6,808, 21,580) |
| **North Dakota** | 11,563 (6,456, 17,318) | 1,268 (685, 1,952) | 300 (163, 460) | 2,054 (1,110, 3,156) | 741 (392, 1,149) |
| **Ohio** | 147,527 (89,066, 210,398) | 20,290 (10,936, 31,025) | 4,964 (2,724, 7,685) | 29,839 (16,551, 45,733) | 14,609 (7,583, 22,993) |
|  | Omicron BA.1* | Omicron BA.2* | Omicron BA.4* | Omicron BA.5* | Omicron XBB* |
| **Oklahoma** | 65,080 (41,002, 90,000) | 4,214 (2,261, 6,481) | 1,683 (917, 2,571) | 11,447 (6,416, 17,522) | 4,384 (2,388, 6,855) |
| **Oregon** | 47,195 (25,717, 69,205) | 12,278 (6,451, 18,790) | 2,193 (1,138, 3,367) | 11,708 (6,036, 18,016) | 3,848 (1,957, 5,961) |
| **Pennsylvania** | 145,978 (78,954, 217,595) | 28,065 (13,808, 45,040) | 4,308 (2,098, 6,965) | 27,038 (13,534, 43,302) | 17,905 (8,334, 29,362) |
| **Rhode Island** | 16,200 (10,789, 21,801) | 3,574 (2,030, 5,372) | 394 (226, 596) | 2,288 (1,321, 3,440) | 2,493 (1,455, 3,710) |
| **South Carolina** | 84,921 (53,210, 115,986) | 7,929 (4,615, 11,681) | 4,136 (2,368, 6,100) | 14,312 (8,188, 21,169) | 4,398 (2,504, 6,496) |
| **South Dakota** | 12,022 (6,712, 18,005) | 1,152 (624, 1,770) | 424 (230, 651) | 2,076 (1,112, 3,203) | 1,385 (733, 2,148) |
| **Tennessee** | 97,662 (59,133, 136,697) | 8,670 (4,949, 12,898) | 3,894 (2,156, 5,847) | 20,986 (11,682, 31,856) | 7,156 (4,035, 10,741) |
| **Texas** | 432,525 (255,142, 619,817) | 39,191 (21,317, 60,325) | 16,480 (8,821, 25,662) | 87,585 (47,036, 134,975) | 28,519 (15,041, 44,741) |
| **Utah** | 42,484 (25,153, 60,734) | 6,060 (3,227, 9,442) | 1,097 (576, 1,710) | 6,908 (3,628, 10,773) | 2,478 (1,301, 3,837) |
| **Vermont** | 6,620 (3,891, 9,483) | 2,692 (1,453, 4,081) | 298 (161, 456) | 1,190 (619, 1,841) | 755 (402, 1,155) |
| **Virginia** | 102,760 (61,246, 143,788) | 19,659 (10,351, 30,465) | 5,019 (2,677, 7,729) | 21,178 (11,449, 32,737) | 11,310 (5,887, 17,766) |
| **Washington** | 91,418 (51,214, 133,457) | 20,114 (10,647, 31,232) | 2,654 (1,382, 4,094) | 18,868 (9,883, 29,248) | 7,000 (3,605, 10,913) |
| **West Virginia** | 20,331 (12,485, 28,457) | 3,877 (2,261, 5,706) | 1,245 (731, 1,832) | 5,565 (3,218, 8,278) | 1,929 (1,084, 2,897) |
| **Wisconsin** | 73,723 (46,292, 102,151) | 13,150 (7,494, 19,581) | 2,316 (1,311, 3,485) | 12,706 (7,374, 19,070) | 6,210 (3,540, 9,358) |
| **Wyoming** | 7,768 (4,382, 11,443) | 1,295 (701, 2,011) | 705 (373, 1,092) | 1,981 (1,028, 3,093) | 642 (327, 1,017) |

**Table S3.**
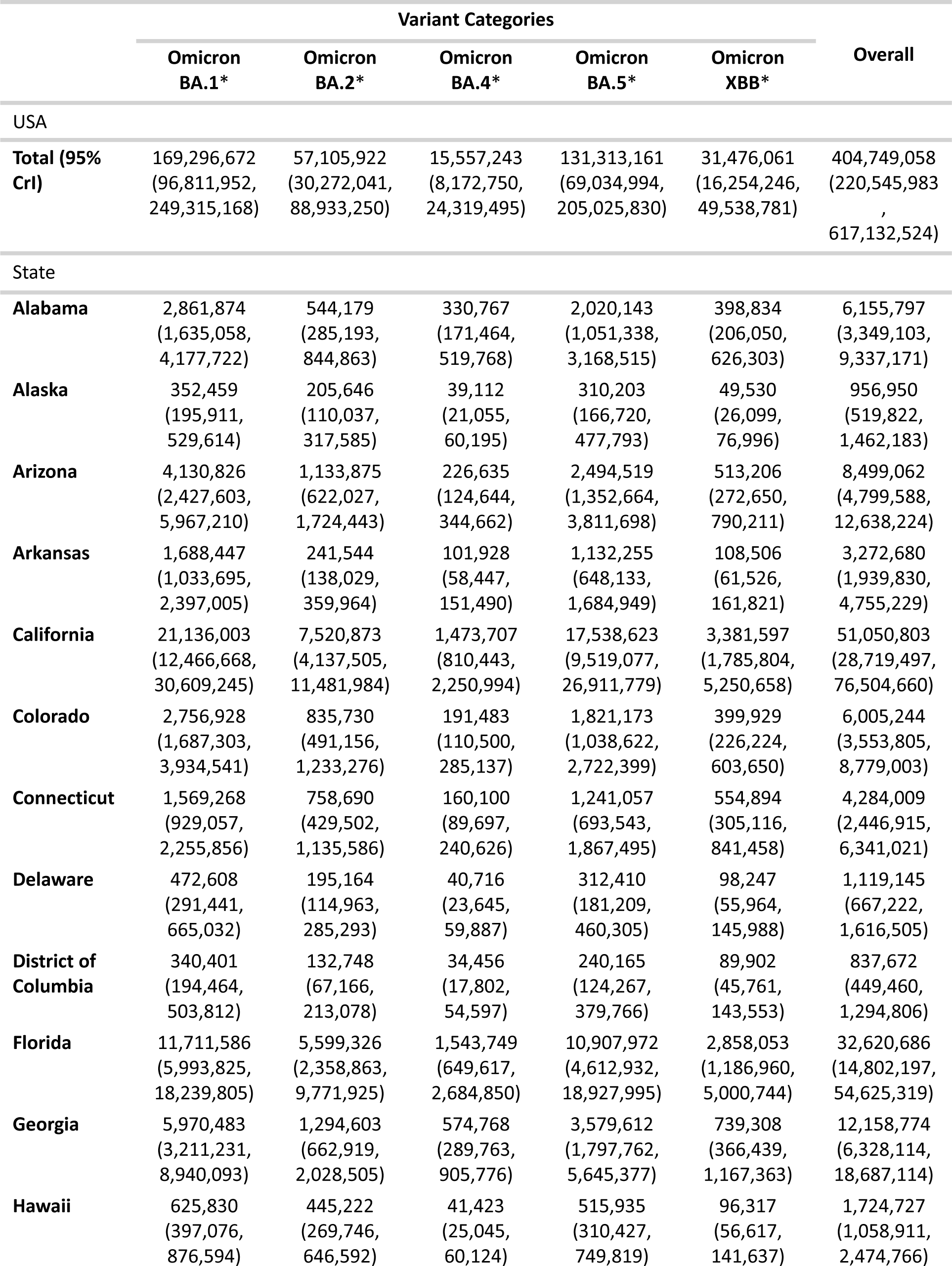
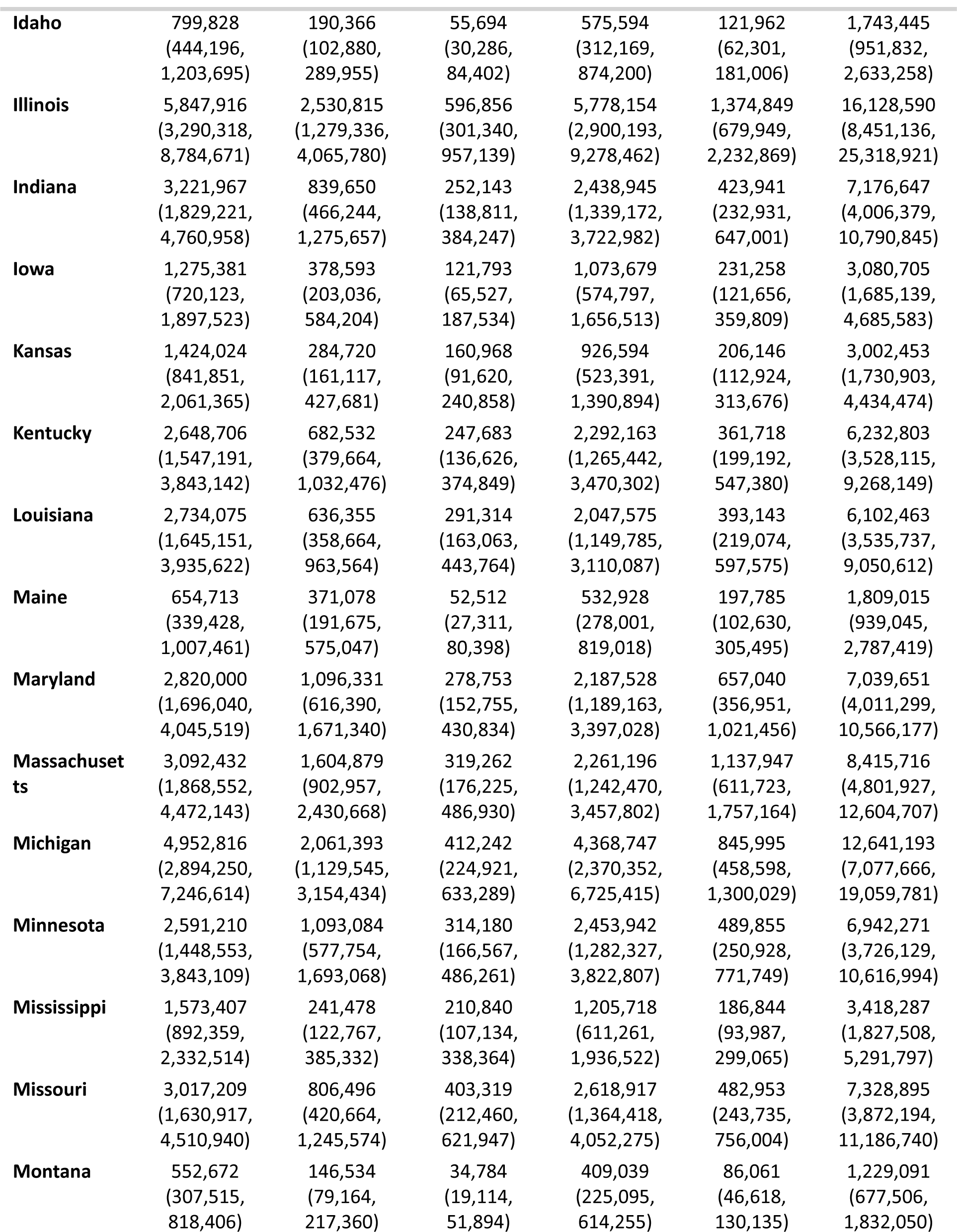
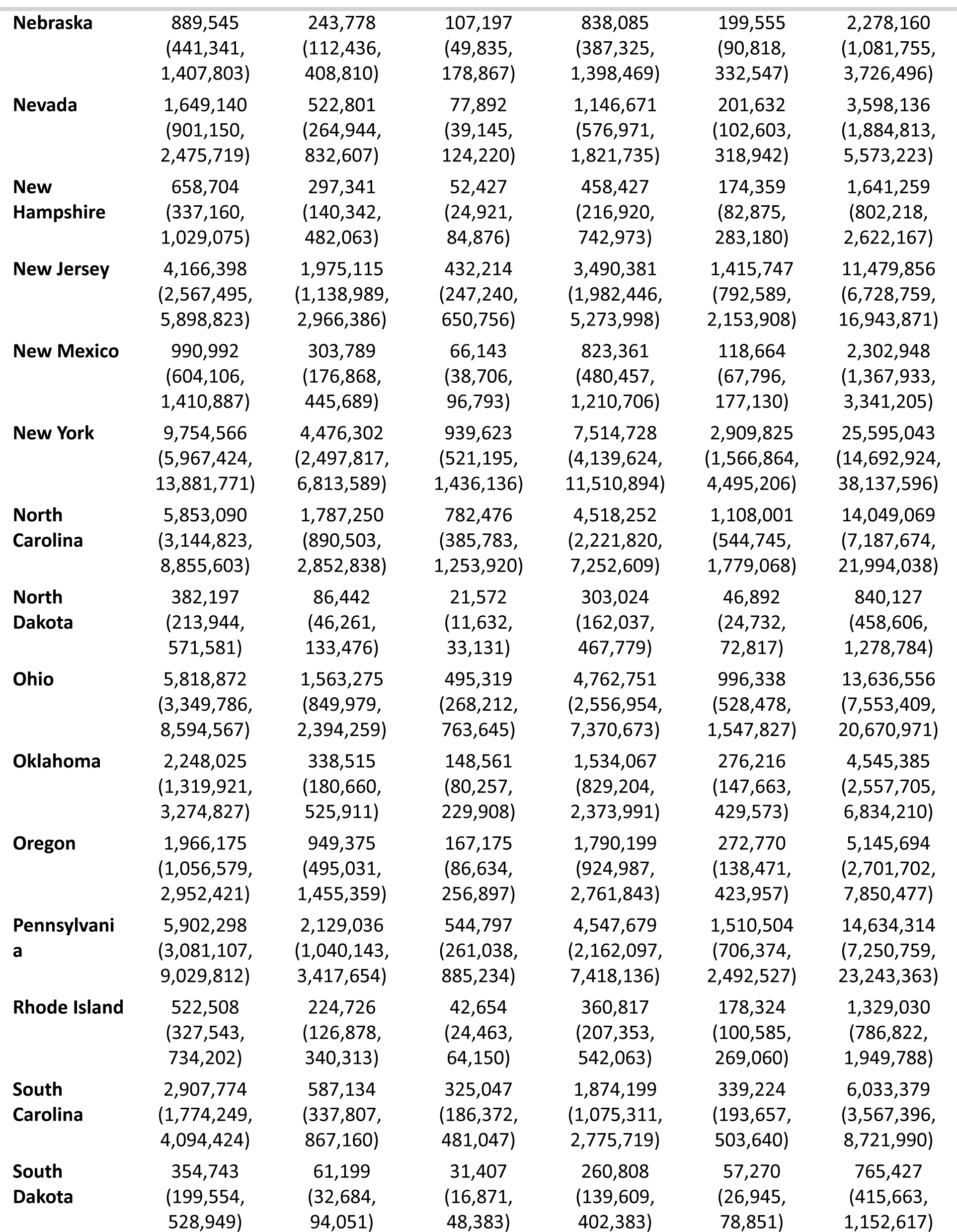
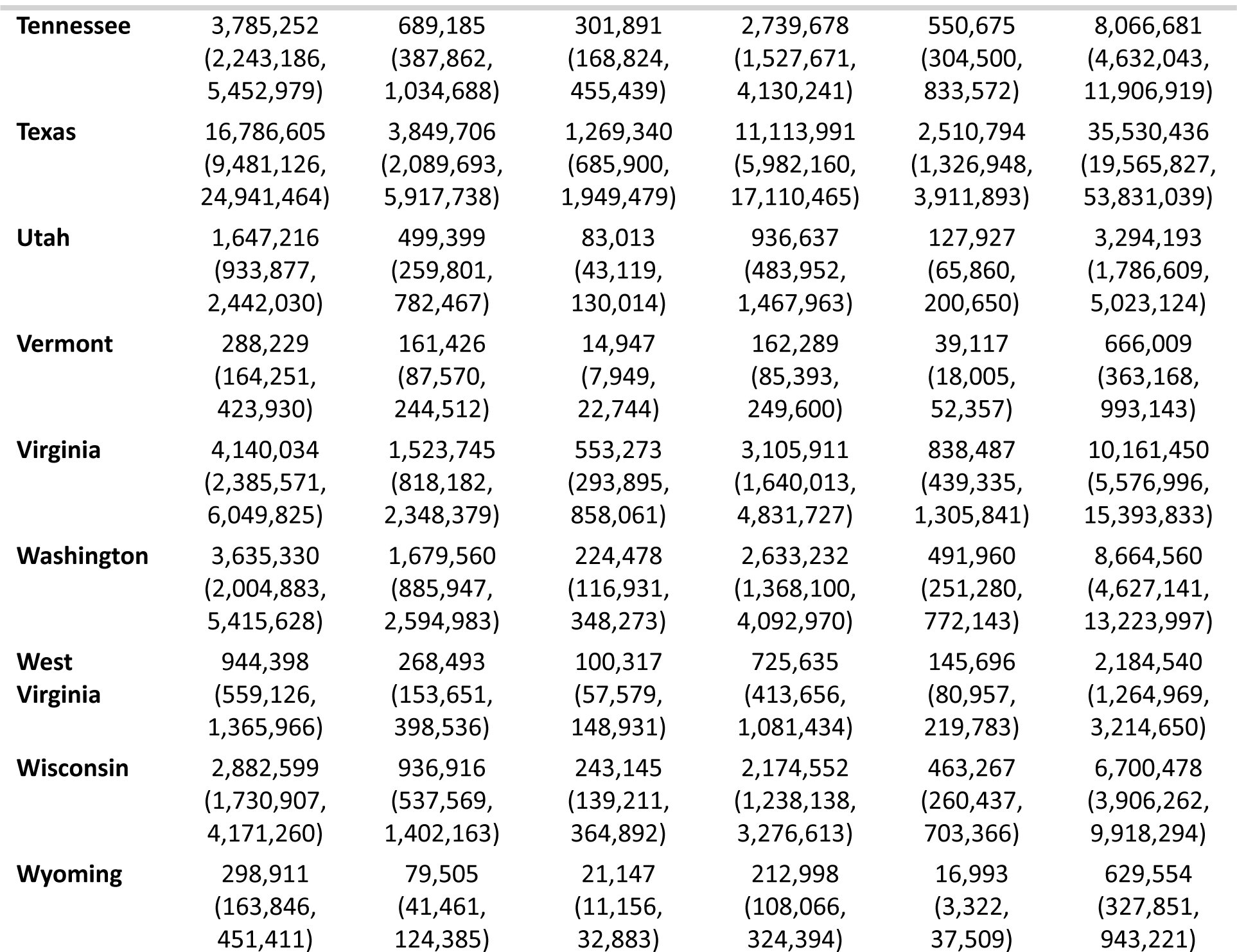
Total of infections by variant categories.

**Table S4.** Attack rates by variant categories.

|  | Variant Categories |  |  |  |  |
| --- | --- | --- | --- | --- | --- |
|  | Omicron BA.1* | Omicron BA.2* | Omicron BA.4* | Omicron BA.5* | Omicron XBB* |
| USA mean (max, min) | 47.7% (57.2%, 37.9%) | 17.2% (29.6%, 6.4%) | 4.4% (6.7%, 2.3%) | 35.7% (47.9%, 24.3%) | 9.1% (15.6%, 3.4%) |
| States |  |  |  |  |  |
| **Alabama** | 56.6% | 10.2% | 6.1% | 37.5% | 7.4% |
| **Alaska** | 45.4% | 25.9% | 4.9% | 39.1% | 6.2% |
| **Arizona** | 54.9% | 14.5% | 2.9% | 31.9% | 6.6% |
| **Arkansas** | 54.2% | 7.5% | 3.2% | 35.2% | 3.4% |
| **California** | 51.5% | 17.6% | 3.4% | 41.0% | 7.9% |
| **Colorado** | 46.0% | 13.6% | 3.1% | 29.6% | 6.4% |
| **Connecticut** | 42.7% | 19.9% | 4.2% | 32.6% | 14.5% |
| **Delaware** | 47.4% | 19.0% | 4.0% | 30.4% | 9.5% |
| **District of Columbia** | 45.8% | 16.7% | 4.4% | 30.7% | 11.4% |
| **Florida** | 50.8% | 21.7% | 6.0% | 42.8% | 11.1% |
| **Georgia** | 54.3% | 11.2% | 5.0% | 31.0% | 6.4% |
| **Hawaii** | 42.6% | 29.6% | 2.8% | 34.4% | 6.4% |
| **Idaho** | 42.1% | 10.0% | 2.9% | 30.2% | 6.9% |
| **Illinois** | 43.2% | 17.7% | 4.2% | 40.7% | 9.6% |
| **Indiana** | 45.7% | 11.5% | 3.5% | 33.5% | 5.8% |
| **Iowa** | 38.3% | 11.0% | 3.6% | 31.4% | 6.7% |
| **Kansas** | 47.0% | 9.1% | 5.2% | 29.7% | 6.6% |
| **Kentucky** | 57.2% | 14.2% | 5.2% | 47.9% | 7.6% |
| **Louisiana** | 56.4% | 12.6% | 5.7% | 40.5% | 7.8% |
| **Maine** | 45.9% | 25.8% | 3.7% | 37.3% | 13.8% |
| **Maryland** | 45.0% | 16.6% | 4.2% | 32.7% | 9.8% |
| **Massachusetts** | 42.6% | 21.5% | 4.3% | 30.2% | 15.2% |
| **Michigan** | 47.2% | 19.0% | 3.8% | 40.2% | 7.8% |
| **Minnesota** | 44.0% | 17.9% | 5.1% | 40.0% | 7.9% |
| **Mississippi** | 50.2% | 7.3% | 6.3% | 35.9% | 5.6% |
| **Missouri** | 47.4% | 12.3% | 6.1% | 39.8% | 7.3% |
| **Montana** | 49.8% | 13.4% | 3.1% | 36.3% | 7.6% |
| **Nebraska** | 42.4% | 10.9% | 4.8% | 37.7% | 9.1% |
| **Nevada** | 51.0% | 15.3% | 2.3% | 33.8% | 6.0% |
| **New Hampshire** | 44.9% | 19.8% | 3.5% | 30.5% | 11.5% |
| **New Jersey** | 45.4% | 20.5% | 4.5% | 36.2% | 14.6% |
| **New Mexico** | 45.7% | 13.8% | 3.0% | 37.1% | 5.3% |
|  | Omicron BA.1* | Omicron BA.2* | Omicron BA.4* | Omicron BA.5* | Omicron XBB* |
| **New York** | 48.4% | 21.2% | 4.4% | 35.4% | 13.7% |
| **North Carolina** | 53.0% | 15.4% | 6.7% | 38.9% | 9.5% |
| **North Dakota** | 47.4% | 10.4% | 2.6% | 36.6% | 5.7% |
| **Ohio** | 47.2% | 12.4% | 3.9% | 37.3% | 7.8% |
| **Oklahoma** | 54.3% | 7.8% | 3.4% | 35.4% | 6.4% |
| **Oregon** | 44.8% | 21.3% | 3.7% | 39.9% | 6.1% |
| **Pennsylvania** | 43.7% | 15.1% | 3.8% | 31.7% | 10.4% |
| **Rhode Island** | 47.7% | 19.5% | 3.7% | 31.4% | 15.6% |
| **South Carolina** | 55.4% | 10.8% | 6.0% | 34.4% | 6.2% |
| **South Dakota** | 37.9% | 6.4% | 3.3% | 27.2% | 7.5% |
| **Tennessee** | 53.6% | 9.4% | 4.1% | 37.5% | 7.5% |
| **Texas** | 55.0% | 12.2% | 4.0% | 35.3% | 7.9% |
| **Utah** | 48.8% | 14.2% | 2.4% | 26.8% | 3.7% |
| **Vermont** | 44.3% | 24.4% | 2.3% | 24.3% | 7.5% |
| **Virginia** | 46.7% | 16.5% | 5.9% | 33.3% | 9.0% |
| **Washington** | 45.8% | 20.5% | 2.7% | 32.0% | 5.9% |
| **West Virginia** | 50.7% | 14.1% | 5.3% | 38.0% | 7.6% |
| **Wisconsin** | 47.2% | 15.0% | 3.9% | 34.5% | 7.3% |
| **Wyoming** | 48.6% | 12.6% | 3.4% | 35.7% | 6.5% |

**Table S5.** Interval values to the R_t_ estimates by variant categories.

|  | Variant Categories |  |  |  |  |
| --- | --- | --- | --- | --- | --- |
|  | Omicron BA.1* | Omicron BA.2* | Omicron BA.4* | Omicron BA.5* | Omicron XBB* |
| max (p25, p75) |  |  |  |  |  |
| **median** | 4.23 (0.80, 1.06) | 3.94 (0.88, 1.14) | 2.59 (0.91, 1.10) | 2.94 (0.92, 1.08) | 2.35 (0.97, 1.20) |
| **upper** | 7.74 (0.89, 1.27) | 6.20 (0.94, 1.22) | 3.97 (0.96, 1.18) | 4.79 (0.96, 1.12) | 3.68 (0.99, 1.32) |
| **lower** | 2.13 (0.70, 0.95) | 2.00 (0.82, 1.06) | 1.65 (0.86, 1.04) | 1.86 (0.88, 1.04) | 1.56 (0.94, 1.10) |

